# ASSESSING THE IMPACT OF MEAL VOLUME ON BODY SURFACE GASTRIC MAPPING METRICS IN HEALTHY CONTROLS

**DOI:** 10.64898/2026.03.19.26348835

**Authors:** India Fitt, Mikaela Law, Gen Johnston, Charlotte Daker, Sam Simmonds, Billy Wu, Nicky Dachs, Gabriel Schamberg, Chris Varghese, Armen Gharibans, Thomas L Abell, Christopher N. Andrews, Greg O’Grady, Stefan Calder

## Abstract

**Background:** Chronic gastroduodenal symptoms are challenging to diagnose and treat. Body surface gastric mapping provides non-invasive biomarkers of gastric function, but the requirement of a standard meal for postprandial assessment can be difficult for severely symptomatic patients.

**Aims:** To assess the impact of reduced meal sizes and fasting on body surface gastric mapping metrics to determine clinical interpretability under non-standard nutritional loads.

**Methods:** Healthy controls (n=60) underwent a 4.5-hour Gastric Alimetry test. Three age, sex, and BMI-matched groups (n=20 each) were compared: Standard Meal (482 kCal), Nutrient bar + Water (250 kcal), and Fasted (no meal). Principal Gastric Frequency, Gastric Alimetry Rhythm Index, BMI-Adjusted Amplitude, and fed:fasted Amplitude Ratio were analyzed against normative intervals.

**Results:** Meal status significantly affected amplitude-based metrics; the Standard Meal group exhibited higher BMI-Adjusted Amplitude (p<0.001) and fed:fasted Amplitude Ratio (p=0.001) than Fasted and Bar + Water groups. Frequency and rhythm-based metrics were resilient; Principal Gastric Frequency (p=0.245) and Gastric Alimetry Rhythm Index (p=0.336) showed no significant differences across conditions. While amplitude deviations were common in the Fasted group (20% fell below the normative range), Gastric Alimetry Rhythm Index and Principal Gastric Frequency remained within normal reference ranges for 95% of participants across all conditions.

**Conclusions:** While consuming <50% of the standard meal significantly reduces gastric amplitude, gastric rhythm remains stable. Principal Gastric Frequency and Gastric Alimetry Rhythm Index function as reliable biomarkers of gastric myoelectrical function regardless of nutritional state.

## Introduction

Chronic gastroduodenal symptoms, such as nausea, bloating, abdominal pain, early satiety, and excessive fullness, are prevalent and challenging to diagnose and treat due to their overlapping and multifactorial nature [1–3]. These symptoms are common in patients with gastric diseases, such as gastroparesis [3, 22], and with gastroduodenal disorders of gut-brain interaction (DGBIs), such as chronic nausea and vomiting syndrome (CNVS) and functional dyspepsia (FD) [2]. These patients often undergo extensive testing and trial-and-error treatment approaches, with little symptom improvement [4]. Accurate assessment of gastric function is essential for identifying underlying pathophysiology and guiding treatment strategies.

Body surface gastric mapping (BSGM) is a non-invasive test that provides detailed insights into gastric bioelectrical activity and function. BSGM measures novel biomarkers that may help differentiate gastroduodenal conditions according to underlying disease mechanisms [10]. A commercially available BSGM test, Gastric Alimetry™ (Alimetry Ltd., New Zealand), combines high-resolution electrogastrography with validated symptom profiling and mental health assessments [5–9].

The standard Gastric Alimetry test provides patients with a standardized test meal, which is used to evaluate the stomach’s postprandial response by providing a physiological challenge for assessing neuromuscular and bioelectrical activity [19]. This standardized meal consists of a widely available nutrient drink and energy bar (totalling 482 kCal), which has been used to establish normative reference intervals for BSGM metrics [13]. However, for patients with severe gastroduodenal symptoms, consuming a full standardized meal can be challenging, potentially limiting the test’s applicability in these populations. Moreover, while the effects of meal size variations on BSGM outcomes have been explored [10], the impact of fasting status on gastric bioelectrical patterns remains under-examined.

This study therefore aimed to evaluate the impact of reduced meal sizes and fasting conditions on BSGM test metrics in healthy controls. By comparing gastric bioelectrical activity across different meal conditions (complete fasting, consumption of the solid portion of the meal only, and the full standardized meal) this study sought to determine whether BSGM reports remain interpretable with smaller meals or fasting.

## Methods

Data were collected in Auckland (New Zealand), Calgary (Canada), and Louisville (KY, USA). Ethical approvals were granted by the Auckland Health Research Ethics Committee (AHREC; AH1130), the University of Calgary Conjoint Health Research Ethics Board (REB19-1925), and the University of Louisville Institutional Review Board (IRB: 20.0962; 737935). All participants provided written informed consent.

### Participants

Participants were recruited into and from a prospective multi-national consortium database (https://www.bsmconsortium.com/) and assigned to one of three groups (n = 20 each): Standard Meal, Bar + Water and Fasted (meals detailed below). Participant groups were age, sex and BMI matched. The Fasted and Bar + Water groups were recruited in New Zealand, while the Standard Meal group was recruited in New Zealand, Calgary (Canada), and Louisville (KY, USA).

Healthy volunteers were recruited via local advertisement (November 2023 - March 2024). Eligible participants were over 18 years old with no history of abdominal surgery and no use of medications affecting gastrointestinal function (prokinetics, antispasmodics, opioid analgesics, or sedatives). Participants were excluded for: active gastrointestinal symptoms or diseases; a history of gastroduodenal disorders meeting Rome IV criteria for CNVS or FD; regular (> 1x/week) cannabis use; BMI over 35 kg/m^2^; active abdominal wounds or abrasions; fragile skin; adhesive allergies; pregnancy; or prior upper GI surgeries, per the Gastric Alimetry Instructions for Use [13]

### Study Protocol

The standard Gastric Alimetry test protocol was followed, as described in previous literature [8–10, 13]. All studies commenced in the morning, with all participants fasting for > 6 hrs and asked to avoid caffeine and nicotine on the day prior to testing. The participants’ abdominal skin was shaved if required, prepared using NuPrep (Weaver & Co, CO, USA), and customized array placement occurred according to patient surface anatomy measurements. Recordings were performed for a fasting period of 30 min, followed by the specified test meal (for the Bar + Water group and Standard Meal group) consumed within 10 min, and a 4-hr postprandial recording to capture a full gastric activity cycle. Participants sat reclined in a chair and were asked to limit movement, talking, and sleeping, but were able to read, watch media, work on a mobile device, and mobilize for comfort breaks.

To ensure consistency in metric calculation, particularly for the Fed:Fasted Amplitude Ratio (ff:AR), the Fasted group logged a ‘meal’ on the Gastric Alimetry App at the standard mealtime. This simulated a clinical scenario where a patient is unable to consume the test meal, allowing us to evaluate the impact on standard reports while controlling for variables like circadian rhythms and the effects of a prolonged seated position. This ensured that the ‘postprandial’ metrics in the Fasted group were calculated identically to the fed groups, isolating the effect of caloric intake.

### Test meals

The Standard Meal group consumed the standard Gastric Alimetry meal; an energy bar (250 kcal, 5 g fat, 45 g carbohydrate, 10 g protein, 7 g fiber; Clif Bar & Company, CA, USA) and Ensure nutrient drink (232 kcal, 250 mL; Abbott Nutrition, IL, USA), or a calorie-matched diabetic or gluten-free alternative. The Bar + Water group consumed the same energy bar and 250 mL of water. Lastly, the Fasted group did not consume a meal during the testing period.

### Outcome Measures

#### Gastric Alimetry Metrics

The Gastric Alimetry system outputs a spectrogram for each examination, defining the frequency and amplitude characteristics of the recorded signals over time [10]. From these spectrograms, four distinct spectral metrics are calculated, as previously described [13–16].

□ *BMI-Adjusted Amplitude (μV):* Measures the overall intensity of the recorded gastric electrical activity, with adjustments made to normalize for Body Mass Index (BMI).
□ *Gastric Alimetry Rhythm Index™ (GA-RI):* Quantifies gastric rhythm stability by assessing the concentration of amplitude within the gastric frequency band over time, providing a value between 0 and 1. This metric is also adjusted for BMI.
□ *Principal Gastric Frequency (PGF) (cpm):* Identifies the dominant frequency associated with stable and persistent gastric electrical activity.
□ *Fed: Fasted Amplitude Ratio (ff:AR):* Assesses the gastric response to a meal stimulus, calculated as the ratio of the maximum 1-hour average postprandial amplitude to the pre-prandial amplitude.

The Principal Gastric Frequency, BMI-Adjusted Amplitude, and GA-RI are considered the primary metrics in BSGM, as they enable patient classification into subgroups based on established normative reference intervals [13, 17]. The ff:AR serves as a supportive metric for assessing potential neuromuscular dysfunction. However, deviations from the normative reference intervals for ff:AR alone do not necessarily indicate abnormality, as this metric can be influenced by other physiological factors, such as migrating motor complexes and diurnal variations [7, 18].

#### Statistical Analysis

IBM SPSS Statistics v30 was used for analysis. A sample size of n=20 per group was utilized, which is consistent with established protocols in comparable BSGM and gastrointestinal electrophysiology studies [7, 9, 12]. One-way ANOVAs were used to compare the four BSGM metrics between the three groups across the whole recording duration and as ‘hour-by-hour’ comparisons to examine temporal changes. Specifically, the groups were compared across the following time-points: baseline (pre-meal for the fed groups), 1–2 hours post-meal, 2–3 hours post-meal, and 3–4 hours post-meal. The ff:AR metric, which is calculated based on the entire 4.5-hour test period, was not included in the hour-by-hour analysis. Variations in degrees of freedom for PGF analysis reflect the exclusion of individual recordings where a dominant frequency could not be definitively isolated. For non-significant rhythm metrics, effect sizes and 95% confidence intervals (CIs) are reported to provide further context on the magnitude of the observed trends [23].

For each study group, the proportion of individuals falling outside the established normative reference intervals [11] was analysed using a one-tailed binomial test with the probability parameter set at 0.05 (expected proportion of individuals outside the reference range).

For all tests, statistical significance was set to *p* < 0.05. Significance thresholds were adjusted for multiple comparisons, when appropriate, using post-hoc comparisons using Bonferroni corrections.

## Results

BSGM using Gastric Alimetry was completed in 60 healthy subjects. These 60 subjects were split into 3 groups (Fasted, Bar + Water and Standard Meal). There were no significant differences between the groups in terms of age, gender, or BMI (**Table 1**).

**Table 1.** Demographic characteristics and BSGM metrics.

| Baseline Variable | Fasted | Bar + Water | Standard Meal | p-value | $\eta^2$ [Confidence Interval] |
| --- | --- | --- | --- | --- | --- |
| Median Age, years (range) | 34.5 (22-62) | 33.0 (18-78) | 33.0 (22-61) | 0.962<sup>a</sup> |  |
| Gender, n (%) |  |  |  |  |  |
| Male | 6 (30) | 5 (25) | 7 (35) | 0.788<sup>b</sup> |  |
| Female | 14 (70) | 15 (75) | 13 (65) |  |  |
| BMI Mean (SD), kg/m<sup>2</sup> | 24.76 (3.38) | 24.72 (2.34) | 24.72 (3.75) | 0.999<sup>a</sup> |  |
| *BSGM Metrics* |  |  |  |  |  |
| PGF, Mean (SD) | 2.97 (0.17) | 3.02 (0.16) | 3.06 (0.19) | 0.245<sup>a</sup> | 0.05 [0.00 - 0.17] |
| BMI-Adjusted Amplitude Mean (SD) | 27.26 (7.46) | 27.61 (7.51) | 37.51 (11.85) | <0.001<sup>a</sup> | 0.22 [0.05 - 0.37] |
| GA-RI, Mean (SD) | 0.49 (0.18) | 0.44 (0.11) | 0.50 (0.15) | 0.336<sup>a</sup> | 0.04 [0.00 - 0.15] |
| ff:AR, Mean (SD) | 1.13 (0.37) | 1.63 (0.74) | 1.82 (0.56) | <0.001<sup>a</sup> | 0.21 [0.04 - 0.37] |

NOTE: BMI = Body Mass Index; SD = Standard Deviation; PGF = Principal Gastric Frequency, ff:AR = Fed-to-fasted Amplitude Ratio; GA-RI = Gastric Alimetry Rhythm Index; <sup>a</sup>= p Value was calculated by one-way ANOVA; <sup>b</sup> = p Value was calculated by $\chi^2$ tests. Asterisks denote significant differences (\* for $p < 0.05$ ; \*\* for $p < 0.01$ ); ns = non-significant.

**Table 2.** Proportion of Participants Outside Normative Reference Intervals for BSGM Metrics.

| BSGM Metric (normative reference range) | Fasted |  | Bar + Water |  | Standard Meal |  | Expected % outside interval |
| --- | --- | --- | --- | --- | --- | --- | --- |
|  | n (%) | p-value | n (%) | p-value | n (%) | p-value |  |
| PGF (2.65cpm - 3.35 cpm) | 0 | - | 0 | - | 0 | - | 5% |
| GARI (>0.25); | 1 (5%) | 0.736 | 1 (5%) | 0.736 | 0 | - | 5% |
| BMI-adjusted amplitude | 4 (20%) | **0.016\*** | 5 (25%) | **0.003\*** | 1 (5%) | 0.736 | 5% |
| ff:AR ($\geq 1.08$); | 11 (55%) | **<0.001\*** | 3 (15%) | 0.075 | 1 (5%) | 0.736 | 5% |

NOTE: BMI = Body Mass Index; SD = Standard Deviation; PGF = Principal Gastric Frequency, ff:AR = Fed-to-fasted Amplitude Ratio; GA-RI = Gastric Alimetry Rhythm Index; <sup>a</sup>= p Value was calculated by one-way ANOVA; <sup>b</sup> = p Value was calculated by $\chi^2$ tests. Asterisks denote significant differences (\* for $p < 0.05$ ; \*\* for $p < 0.01$ ); ns = non-significant.

### Comparison of BSGM Metrics between groups

There were significant differences between the three groups in the overall BMI-Adjusted Amplitude (F(2, 57) = 8.048, p <.001, η^2^ = 0.22). Post hoc comparisons revealed that the Standard Meal group exhibited a significantly higher amplitude (see **Table 1** and **Figure 1**) compared to both the Fasted group (p = 0.002) and the Bar + Water group (p = 0.004).

**Figure 1.**
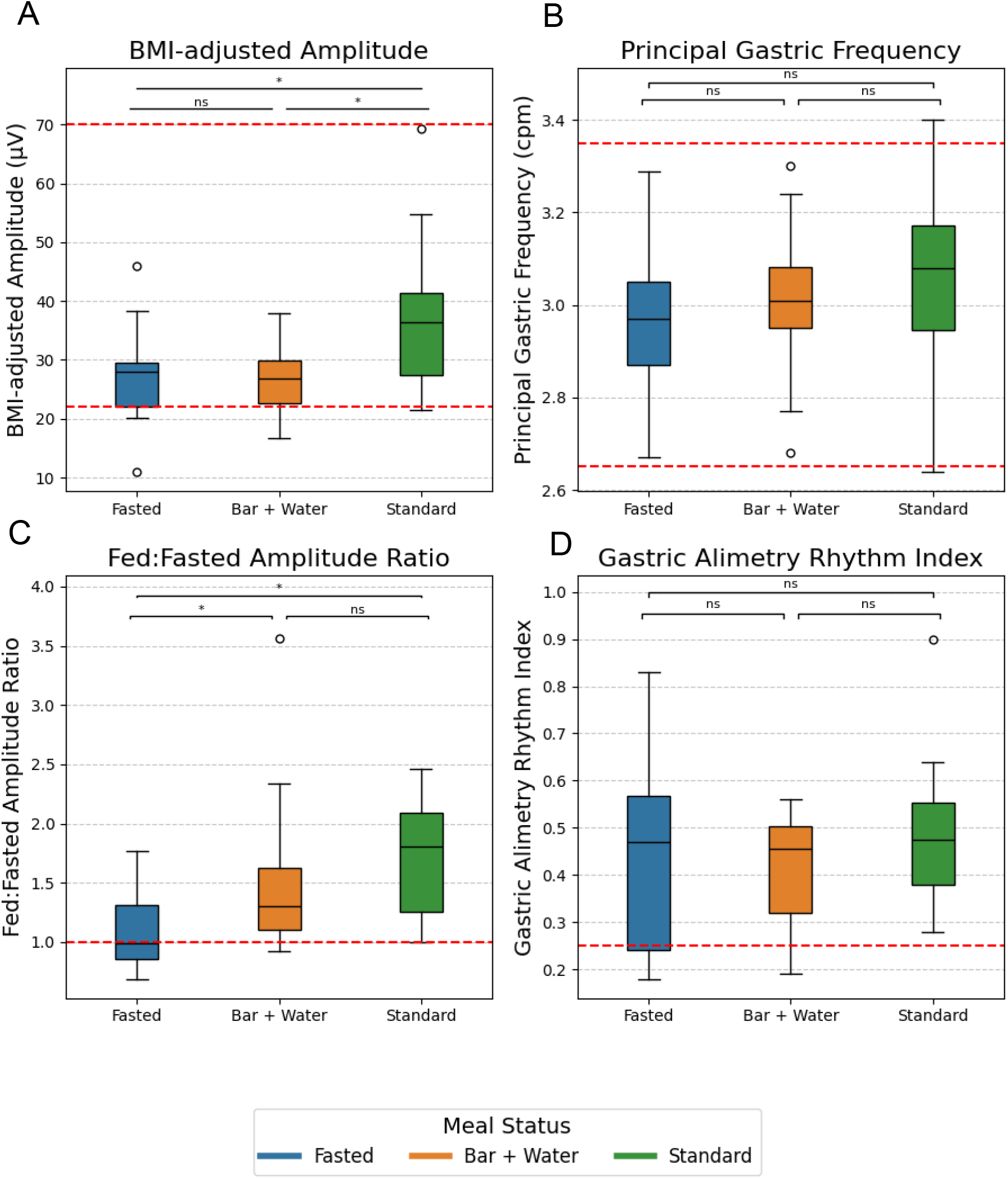
Relationship between meal status (Fasted, Bar + Water and Standard Meal) and key physiological metrics. Box-and-whisker plots illustrating the distribution of spectral metrics across the three experimental groups (Fasted, Bar + Water, and Standard Meal). (A) BMI-adjusted Amplitude (uV): The Standard Meal group exhibited significantly higher overall intensity compared to both the Fasted (p = 0.002) and Bar + Water groups (p = 0.004). (B) Principal Gastric Frequency (cpm): No significant differences were observed across conditions (p = 0.245). (C) Fed:Fasted Amplitude Ratio (ff:AR): The Standard Meal group showed the highest meal response, significantly exceeding the Fasted group (p = 0.001). (D) Gastric Alimetry Rhythm Index (GA-RI): Rhythm stability remained consistent across all meal conditions (p = 0.336). Red dashed lines indicate established normative reference intervals. Asterisks denote significant differences (* for p < 0.05; ** for p < 0.01); ns = non-significant.

The one-way ANOVA for ff:AR also showed significant differences between groups (F(2, 57) = 7.625, p = 0.001, η^2^ = 0.21). As shown in **Table 1** and **Figure 1**, the Standard Meal group showed significantly higher ff:AR than the Fasted group (p = 0.001). Additionally, a significant difference was observed between the Fasted group and the Bar + Water group (p = 0.025).

In contrast, GA-RI (F(2, 57) = 1.111, p =.336, η^2^ = 0.04) and Frequency (F(2, 55) = 1.443, p = 0.245, η^2^ = 0.05) did not demonstrate significant differences across meal conditions.

### Hour-by-hour comparisons

Examination of the time-course data in **Figure 2** suggests that the group-level differences in BMI-adjusted amplitude were primarily determined by the post-prandial rise observed following the Standard Meal. While the Fasted and Bar + Water conditions maintained relatively constant amplitude levels throughout the study, the Standard Meal group demonstrated a significant increase that was detectable by the 1st hour (p = 0.037) and continued through the 4th hour (see **Table 3)**. The effect sizes from the 2nd hour onwards were notably large (η^2^ ≥ 0.21), indicating a substantial physiological response that was absent in the other two conditions. Consistent with expectations, baseline amplitudes, recorded while all protocols remained identical, were statistically indistinguishable across all three groups (p = 0.224)

**Table 3.**
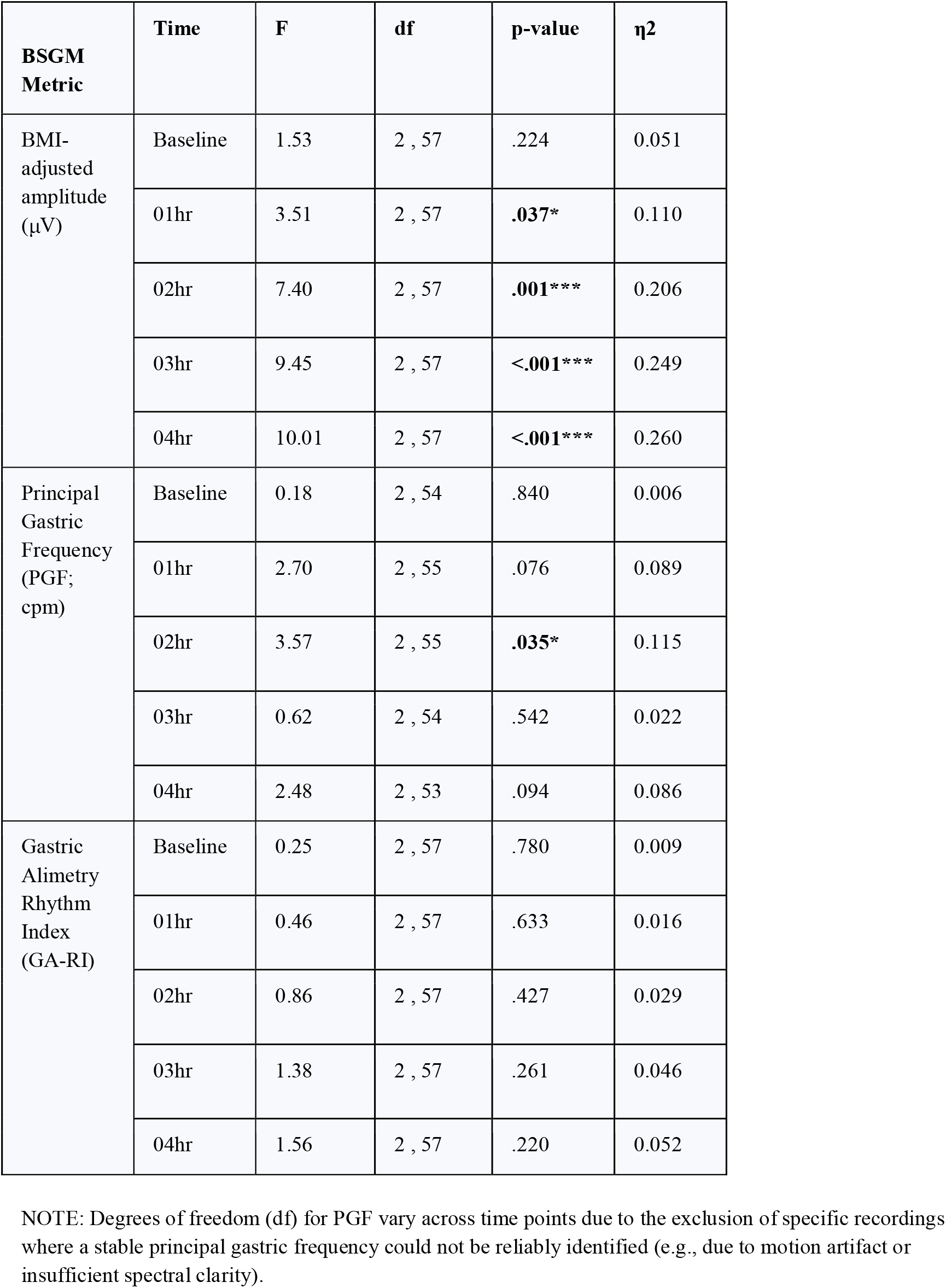
Statistical summary of group differences across time.

**Figure 2.**
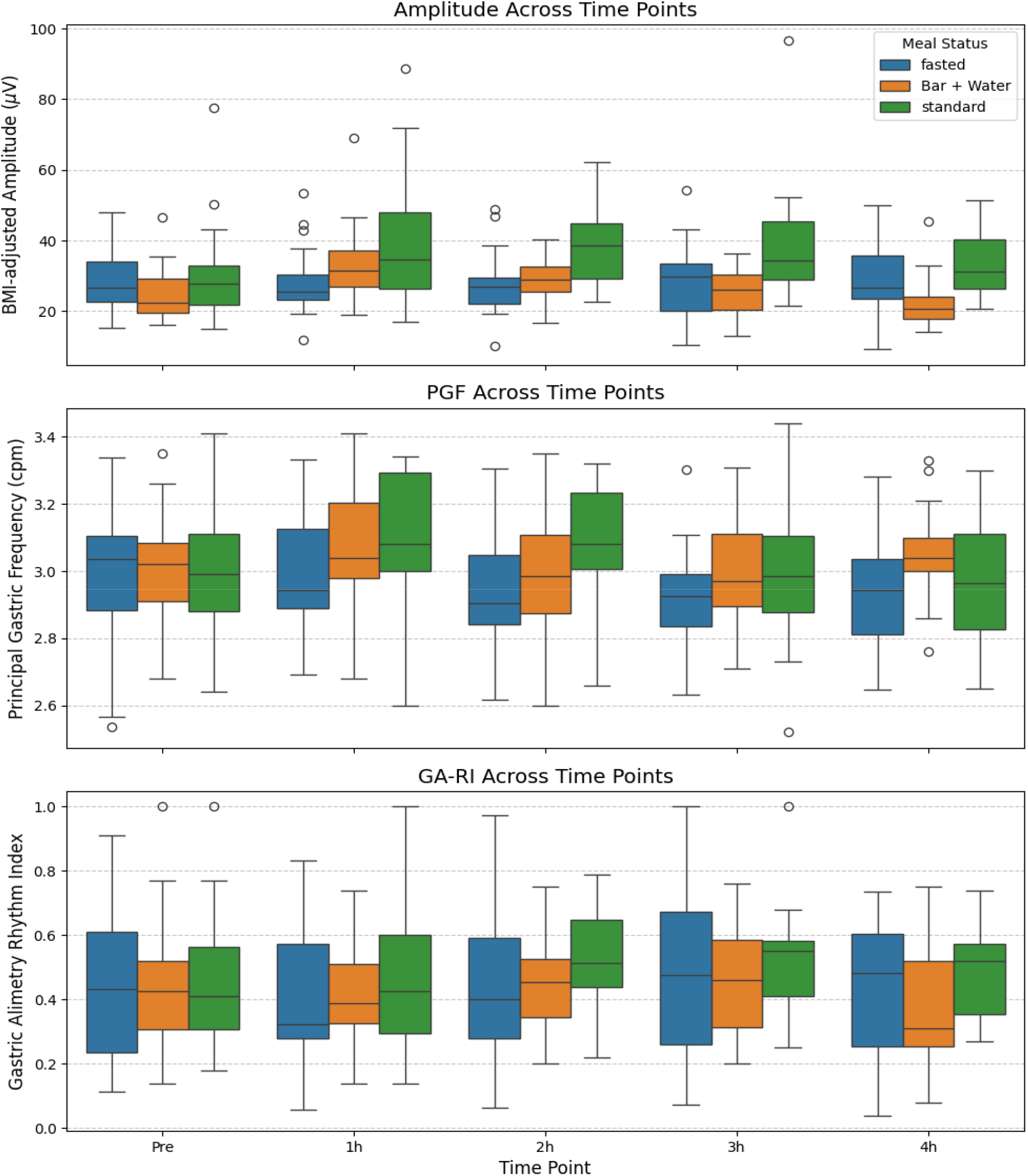
Temporal stability of BSGM metrics across pre- and post-prandial intervals. Time-course analysis of gastric bioelectrical activity at baseline (Pre) and hourly intervals following the meal stimulus. (Top) BMI-adjusted Amplitude (uV): Baseline values were indistinguishable (p = 0.224), but the Standard Meal group demonstrated a significant post-prandial rise starting at Hour 1 (p = 0.037). (Middle) Principal Gastric Frequency (PGF; cpm): A transient shift occurred at Hour 2 in the Standard Meal group (p = 0.035), though group means remained within the normative range (2.65 - 3.35 cpm). (Bottom) Gastric Alimetry Rhythm Index (GA-RI): Gastric rhythm showed minimal fluctuation and no significant differences between groups at any time point (p ≥ 0.220). Box plots represent the median and interquartile range; outliers are indicated by open circles.

For gastric frequency (PGF/cpm), the Standard Meal condition induced a temporary shift at the 2-hour mark, representing the only point of statistical difference between the three groups (F(2, 55) = 3.57, p = 0.035, η^2^ = 0.12). At this interval, mean frequencies for the Standard Meal group (3.15 [± 0.31] cpm) were slightly higher than those observed in the Fasted group (2.96 [± 0.20] cpm) and Bar + Water group (3.00 [± 0.19] cpm) (See supplementary data). While this difference reached statistical significance, all group means remained within the normal reference range (2.65–3.35 cpm); therefore, the observed variance is likely not clinically meaningful. This shift appeared to be short-lived, as group values converged again by the 3rd hour (p = 0.542), likely reflecting the peak of the postprandial response in the fed groups. No significant frequency shifts were detected either at baseline or during the initial hour after the meal.

Finally, visual analysis of GA-RI in **Figure 2** shows minimal fluctuations across both time and condition. This observation is confirmed by the statistical analysis, which yielded no significant group differences at any time point (p ≥ 0.220), with negligible effect sizes recorded throughout the 4-hour post-prandial window (η^2^ ≥ 0.05).

### Comparison to Normative values

Normative data are constructed as 95% intervals, such that 5% of healthy controls are expected to fall outside interval bounds. In this study, observed proportions varied by metric and meal condition (**Table 3**).

The Standard Meal group aligned with these expectations across all metrics (p = 0.736). However, there were more pronounced differences in the other conditions. The Fasted group exhibited significantly higher proportions outside the reference range for ff:AR (55%, p < 0.001); though this was to be expected as there was no meal to respond to. The Fasted group also had 20% (p =.016) outside of normal BMI-adjusted amplitude range. Similarly, the Bar + Water group showed a significant deviation for BMI-adjusted amplitude (25%, p = 0.003).

Across all conditions, GA-RI and PGF remained within normative intervals (p ≥ 0.736) indicating that frequency and rhythm-based markers are resilient to variations in a reduced caloric intake.

## Discussion

This study demonstrates that meal volume and composition primarily modulate gastric myoelectrical amplitude metrics (BMI-Adjusted Amplitude and ff-AR), while the fundamental gastric frequency and rhythm metrics (PGF and GA-RI) remains stable across varying nutritional loads. Although the Standard Meal elicited the most robust physiological increase in gastric amplitude, the Bar + Water protocol (250 kcal) was sufficient to trigger a response and maintain diagnostic integrity for frequency and rhythm biomarkers within established normative reference intervals [13]. This evidence suggests that a reduced caloric protocol may provide a viable testing alternative for patients with severe meal intolerance, ensuring the Gastric Alimetry test remains clinically useful even when a full meal cannot be consumed.

The ability of the test to tolerate variability in the meal makes it highly appropriate for use where patients may have a wide range of nutrient sensitivities, dietary philosophies, severe symptoms related to intake, or may be tube fed. This contrasts with gastric emptying scintigraphy, where slight deviations from the standard egg-based meal may adversely affect interpretability. These results also provide a practical framework for interpreting Gastric Alimetry tests in patients with severe symptoms. While the Standard Meal remains the gold standard for a full physiological challenge, the Bar + Water protocol is a viable clinical alternative for patients with significant early satiety, as it maintains diagnostic integrity for key rhythm markers. Specifically, GA-RI and PGF, and their associated phenotypes such as dysrhythmia or high-frequency activity, can be reliably assessed even when an incomplete meal is consumed. However, clinicians should exercise caution when evaluating phenotypes related to the strength or timing of the meal response (e.g., low meal response, sensorimotor patterns, or delayed onset symptoms). Since BSGM normative reference ranges were established using full meal stimuli [13], clinicians should consider low or borderline amplitudes in fasted or low-volume tests as potentially reflective of reduced caloric intake rather than evidence of neuromuscular or myoelectrical impairment [24]. A key finding of this research is the high reliability and resilience of rhythm-based metrics across varying prandial states. PGF remained stable across all meal groups (2.65 cpm - 3.35 cpm), with no significant differences observed between the fasted, low-volume, and standard meal groups. Similarly, GA-RI showed minimal fluctuations across both time and meal conditions, yielding negligible effect sizes throughout the 4-hour post-prandial window. Because GA-RI and PGF consistently remained within expected normative limits regardless of meal status, these metrics can be considered the most “resilient” biomarkers of the underlying gastric myoelectrical activity.

In contrast to the stability of the rhythm, amplitude-based metrics such as BMI-Adjusted Amplitude and ff:AR were highly sensitive to meal volume [20, 21]. The Standard Meal group exhibited a significantly higher amplitude compared to both the Fasted and Bar + Water groups, driven by the post-prandial rise that was largely absent in the other conditions.

The variability observed during fasting may reflect diurnal fluctuations in gastric amplitude or could potentially represent detection of the underlying Migrating Motor Complex (MMC), which manifests as fluctuations [18]. Current literature lacks a detailed characterization of how these specific MMC phases manifest in BSGM recordings within the fasted stomach [18], representing a key area for future physiological validation against reference standards such as antroduodenal manometry.

While this study is the first to systematically characterize the fasted state using the Gastric Alimetry system, several limitations must be acknowledged. The sample size (n=20 per group) is limited, although it is consistent with established protocols in comparable BSGM and gastrointestinal electrophysiology studies, and was sufficient to show clear statistical outcomes [7, 9, 12]. The study was also conducted in healthy controls only; symptomatic patients with gastric dysrhythmias [22] or functional dyspepsia may exhibit different levels of rhythm stability under stress or fasting conditions, and these results may not be directly generalizable to those populations. Furthermore, although the groups were matched, they were recruited across different international sites; however, the use of a highly standardized protocol across all centres minimizes the impact of geographic variability.

In conclusion, this study establishes that gastric rhythm biomarkers (PGF and GA-RI) are highly resilient to variations in prandial state, providing stable diagnostic metrics regardless of meal volume. While the standard 482 kcal meal remains optimal for evaluating gastric amplitude and post-prandial power shifts, a reduced-caloric protocol (250 kcal) is a valid alternative for patients with severe meal intolerance. Clinicians should interpret amplitude-based phenotypes with caution when a full meal is not consumed but can rely on rhythm-based metrics to identify underlying gastric dysrhythmias. These findings broaden the clinical utility of Gastric Alimetry, ensuring robust patient phenotyping across a wider spectrum of symptom severity.

## Data Availability

All data produced in the present work are contained in the manuscript

